# The psychological effects of quarantine during COVID-19 outbreak: Sentiment analysis of social media data

**DOI:** 10.1101/2020.06.25.20140426

**Authors:** Weisheng Lu, Liang Yuan, Jinying Xu, Fan Xue, Bin Zhao, Chris Webster

## Abstract

We rely on social distancing measures such as quarantine and isolation to contain the COVID-19. However, the negative psychological effects of these measures are non-negligible. To supplement previous research on psychological effects after quarantine, this research will investigate the effects of quarantine amid COVID-19. We adopt a sentiment analysis approach to analyze the psychological state changes of 1,278 quarantined persons’ 214,874 tweets over four weeks spanning the period before, during, and after quarantine. We formed a control group of 1,278 unquarantined persons with 250,198 tweets. The tweets of both groups are analyzed by matching with a lexicon to measure the anxious depression level changes over time. We discovered a clear pattern of psychological changes for quarantined persons. Anxious depression levels significantly increased as quarantine starts, but gradually diminished as it progresses. However, anxious depression levels resurged after 14 days’ quarantine. It was found that quarantine has a negative impact on mental health of quarantined and unquarantined people. Whilst quarantine is deemed necessary, proper interventions such as emotion management should be introduced to mitigate its adverse psychological impacts.

## Introduction

The 2019 novel coronavirus (COVID-19) epidemic broke out in early 2020 in Wuhan, Hubei Province, spreading rapidly to other mainland Chinese cities and then the world. According to various sources, by early June 2020, total confirmed cases of infection had reached 7 million and the death toll 400,000. The US, Brazil, Russia, UK, India, Spain, Italy, France, Germany, China, and Iran are among the hardest hit in terms of confirmed cases. Although reported to have plateaued in some countries, the ascending trend of COVID-19 cases worldwide continues.

Without a vaccine, drug therapies, or other aggressive treatments, we continue to rely heavily on social distancing to slow the spread of COVID-19, or “flatten the curve”.^1^ Social distancing measures have been used for centuries to arrest the spread of infectious disease, and we have seen the closure of schools and workplaces, cancellation of mass events, travel restrictions, voluntary self-isolation, mandatory quarantine, and lockdown of entire communities. Among them, quarantine and isolation are widely exercised during this COVID-19 outbreak.

“Quarantine” refers to the separation and restriction of movement of people who have potentially been exposed to a contagious disease to ascertain whether they have been infected, so reducing their risk of infecting others.^2^ “Isolation” refers to the physical separation of those who have been diagnosed with a contagious disease from those who have not. These terms are often used interchangeably, especially in communications with the public,^3^ and are used interchangeably in this paper, without differentiating whether the grounded subject is infected or not by the COVID-19 virus.

Quarantine is an effective means of pandemic containment. However, placing people in quarantine may have considerable, wide-ranging, and perhaps long-lasting psychological impacts upon them, including fear, anxiety, sadness, depression, grief, and confusion.^2^ Previous studies of quarantine have investigated its psychological effects after the outbreaks of Severe Acute Respiratory Syndrome (SARS) in 2003 and the Middle East Respiratory Syndrome (MERS) in 2012. For example, Hawryluck et al.^5^ examined the psychological effects of quarantine on 129 people in Toronto, Canada, observing symptoms of post-traumatic stress disorder (PTSD) and depression in 28.9% and 31.2% of subjects, respectively. Liu et al.^6^ examined post-outbreak levels of depressive symptoms among hospital employees (*N=549*) exposed to SARS in Beijing 2003 and found that having been quarantined during the outbreak increased the odds of having a high level of depressive symptoms.

Previous studies on the psychological effects of quarantine tend to adopt psychological measurement scales such as IES-R (Impact of Event Scale - Revised) to assess post-traumatic stress^7^; the CES-D (Center for Epidemiologic Studies Depression Scale) to assess depression^6^; GAD-7 (Generalized Anxiety Disorder Questionnaire) to assess anxiety^8^; STAXI-2 (State-Trait Anger Expression Inventory-2) to assess anger^9^; MBI-GS (Maslach Burnout Inventory - General Survey)to assess burnout^10^; and the K10 (Kessler Psychological Distress Scale). to assess psychological distress^11^. These scales are largely subjective and passive sources of measuring psychological effects and are limited by availability, timeliness, and volume of data^12^. Moreover, most of the previous studies only focused on the psychological effect after a pandemic or other traumatic event.

Millions of people around the world remain in lockdown amid the current pandemic, little research has probed into the psychological effects of quarantine during the COVID-19 outbreak. We attribute this lack of empirical research to: (1) insufficient attention paid to the psychological impacts of COVID-19 quarantine; and (2) difficulty to access the data on quarantined people during the ongoing outbreak. Previously, researchers have relied on questionnaire surveys or interviews to understand psychological effects after quarantine. In this paper, we aim to report psychological state changes of quarantine persons before, during, and after quarantine by adopting a novel sentiment analysis approach.

### Sentiment analysis of social media data

People in quarantine have more time to access social media, e.g., Twitter, Facebook, and WeChat. Reports have noted that “during quarantine, iPhone screen time reports are through the roof”^13^. Social media provides a means of self-expression and facilitates measurement of the psychological status of those sharing their feelings which are hard to articulate in traditional means^14^. The huge amount of people’s experience, opinion, and emotion on social media provides a great opportunity for automatic mining and analysis of psychological dynamics^15^.

Sentiment analysis of social media data can enable general large-scale analyses of public mental health status^16^. For example, De Choudhury et al.^17^ used shared Facebook data to characterize and predict postpartum depression among 165 new mothers. Compared to Facebook, Twitter attracts more researchers by an opener data access to the public. For example, Pedersen^18^ screened depression and PTSD among 327 Twitter users; De Choudhury et al.^19^ measured depression among 489 Twitter users. Going beyond traditional methods such as questionnaire survey or interviews, sentiment analysis can help probe into the mental health status of those in quarantine.

### Psychological effects of quarantine as reflected in social media

We focus predominantly on anxious depression—anxiety in depressive individuals— as the most common among mental disturbances in quarantined patients^20,21^. Unlike depression or anxiety alone, which are two independent mental disorders^22^, anxious depression is a mixed anxiety and depression disorder. Depression is characterized by sadness, loss of interest or pleasure in life, feelings of guilt or low self-worth, disturbed sleep or appetite, feelings of tiredness and poor concentration^23^. Anxiety is a conscious state of worry over a future unwanted event, or fear of an actual situation^24^. In this study, we adapted a prevailing sentiment analysis approach of using bag-of-words, to assume the psychological states, e.g., anxious depression, are reflected in various lexicons. Particularly, we developed our lexicon from two sources: (1) the online word lists provided by *MyVocabulary* (https://myvocabulary.com/), a free and interactive vocabulary resource used in over 40,000 schools, and (2) word lists from the scholarly literature^14,19,25^. In developing our lexicon, we integrated words from the two sources and used manual screening to exclude irrelevant or ambiguous terms based on the features of the COVID-19. Our final lexicon comprised 317 words related to anxious depression (see Appendix).

Social media is an unintendedly left big data hub for studying the psychological status of people ^26^. Although a huge amount of social media data is publicly available, deep qualitative investigation into the contents raises ethical concerns over privacy, anonymity and confidentiality, authenticity, voluntary participation and informed consent, data security and management, and sampling, among others^26^. Specific ethical issues differ with the research question, scope of data collection, and whether the aimed analysis is based on quantitative analysis of aggregates of users or qualitative, in-depth examination of individual users^27^. Twitter is a widely used social media platform with API for academic research. The content posted by users on Twitter is by default public^27^, which means that academic researchers are free to access the data. In investigating the psychological impacts of COVID-19 quarantine, this research will undertake quantitative analysis of aggregates of Twitter users, thereby avoiding infringement of Twitter users’ privacy. In addition, we have obtained ethical approval from the University of Hong Kong for this study.

## Methods

We conducted three rounds of data collection and cleansing. The first round aimed to identify persons who had completed a 14-day quarantine period. To start, we read many tweets and interestingly found some typical phrases in use among these persons. We identified quarantined users by searching for tweets containing: 1) “day X in quarantine” and the hashtag “#quarantinelife”, 2) “day X” and the hashtag “#selfquarantine”, and 3) “isolation day X” and the hashtag “#selfisolating”. Here, “X” refers to a number between 1 and 14. Secondly, using the queries, we collected 2,462 users from the Twitter website using the Python tool “TweetScraper”. Thirdly, we used the Natural Language Processing techniques, especially the Text Analysis techniques in Matlab2019a, to process the tweets posted by the potential users and extracted 1,403 quarantined Twitter users. Given our aim to investigate anxious depression levels in individuals before, during, and after quarantine, we also identified individual quarantine starting dates (*D*_*s*_) by reading their tweets. For example, a user’s tweet “self-isolation diary, day 2” dated “2020-03-14” indicates a quarantine start date of 13 March 2020. Lastly, we set the duration of both “after” and “before” quarantine as 7 days to locate the latest quarantine starting date of qualified users as we aimed to collect the 28-day data completely when we extracted the data. After excluding the users whose starting dates were later than the latest quarantine starting dates, we finally obtained 1,278 qualified individuals.

A second-round data collection and processing were conducted to collect the 1,278 qualified users’ tweets and then classify them into one of the setting periods (before, during, and after quarantine). We first used the Python tool to collect all tweets posted by these users in the three periods. After cleansing incorrect or repetitive data, we obtained 214,874 tweets. We then standardized the dates into the three periods, i.e., before quarantine (7 days), during quarantine (14 days), and after quarantine (7 days). It is necessary to adopt a relative date (*D*_*r*_) to replace the absolute date (*D*_*a*_) of each tweet so that we can put all tweets under the three periods for investigation, even though their absolute dates are different. Equation (1) shows the calculation method for the relative date of each tweet.

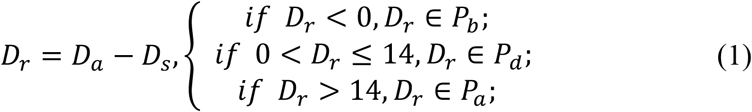

where *P*_*b*_ represents the period 7-days-before-quarantine, *P*_*d*_ represents the period 14-days-during-quarantine, and *P*_*a*_ represents the period 7-days-after-quarantine. After calculating the relative date of 214,874 tweets and classifying them, we obtained 43,341 tweets in the 7 days before quarantine, 120,734 tweets in the 14 days during quarantine, and 50,799 tweets in the 7 days after quarantine.

A third-round data collection and processing were undertaken to collect 1,278 unquarantined users to form a control group. Since the absolute time of 1,278 quarantined users covers from February 15 to April 15, we need to keep the number of quarantined users and unquarantined users consistent in the same absolute date period as the 1,278 quarantined users. We first collected 3,389 users’ tweets posted between February 15 and April 15 2020 (6,204,632 tweets). Afterwards, we defined an unquarantined users as those whose tweets did not contain the hashtag “#isolationlife”, “#selfisolating”, “#selfquarantine”, “#quarantinelife”, “#quarantinequotes”, or the keyword “quarantine” during the sampled period. We thereby obtained 1,498 qualified users. We calculated the number of quarantined users of different periods, and then we matched the distribution of unquarantined user to keep them the same. For example, 100 quarantined users’ 28-day relative dates were from February 15 to March 13 2020, therefore, we distributed 100 unquarantined users into this absolute date period. By this step, we obtained 1,278 unquarantined users from the 1,498 identified ones to be the control group, and two groups have the same user number distribution.

Combining tweet text, post time, and relative date, we obtained a quarantined user data matrix containing 214,874 × 3 data points and an unquarantined user data matrix containing 250,198 × 3 data points.

We developed Equation (2) to quantify the anxious depression (AD) level on Day *i* of all quarantined users:

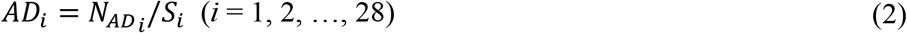

where 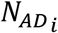 is the number of tweets containing any of the anxious depression keywords (Table S1) on Day *i*; *S*_*i*_ is the total tweet numbers by all quarantined users on Day *i*. For the calculation, we first stripped all tweets texts of each relative date into many single words and calculated the tweet numbers when any of the keywords is hit. The ratios of daily tweet numbers with the keywords to total daily tweet numbers were then calculated to represent the AD level. The top six keywords from the lexicons appearing in the daily tweets were also identified.

### Data analyses, results, and findings

#### General descriptions of the data

We analyzed the tweets posted by the 1,278 qualified individuals before, during, and after quarantine and their 1,278 unquarantined counterparts in the same period. As shown in Figure 1, in general, unquarantined users tweeted more than the quarantined users on average and in total. Only on the first day of quarantine did the quarantined users post more tweets than the unquarantined group. For the quarantined group, daily tweet numbers almost doubled upon starting quarantine. Afterward, their tweet numbers decreased but remained at a high level until one week during quarantine. Tweet numbers then decreased in the second week of quarantine. Given the possibility that the total tweet numbers are attributable to the numbers of users who tweeted, we calculated the average tweet number per quarantined user. As Figure 1 shows, the average tweet number per quarantined user is smaller than that for unquarantined users. Perhaps quarantined users were less willing to express their concerns publicly. Within the quarantined group, the daily tweet frequency increased dramatically when the quarantine starts. The average tweet numbers from Days 1 to 17 (three days after the two-week quarantine) stayed at a relatively higher level than other days.

**Figure 1:**
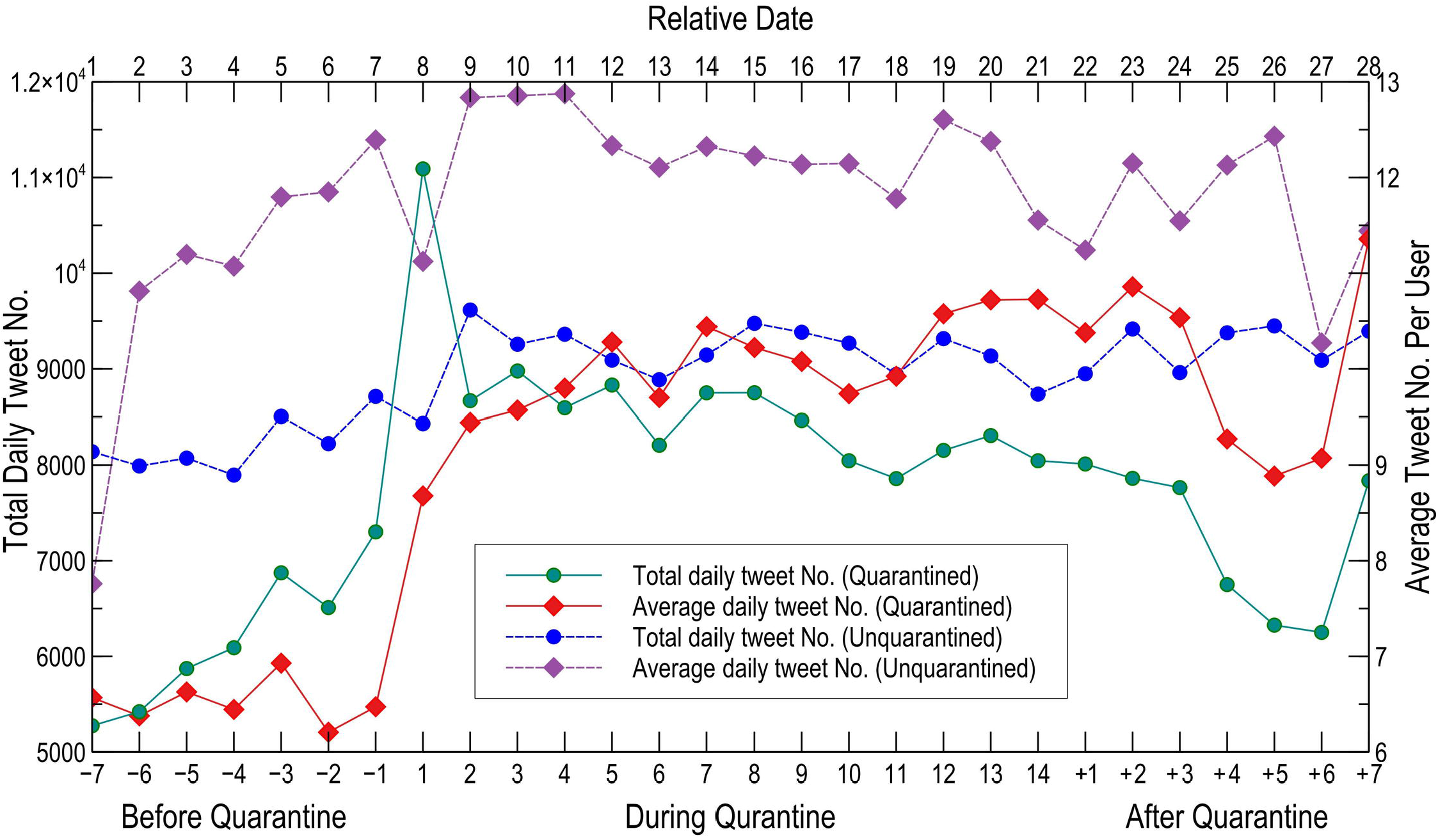
Daily total and average numbers of tweets posted by quarantined users *(N=1,278)*. Tweets were posted by quarantined users whose quarantine finished at least 7 days before data collection. For quarantined users: “Before Quarantine” (Days -7 to -1) is the period before users started their quarantine; “During Quarantine” (Days 1 to 14) is the two-week quarantine period; and “After Quarantine” (Days +1 to +7) is the week after users finished quarantine.

In Figure 1, Total daily tweet numbers of all quarantined users are summarized and displayed (green) on the timeline, and average daily tweet numbers per quarantined user are calculated and displayed (pink). The curves show the changes in number of tweets for the 28 days before, during, and after quarantine. For the unquarantined users, numbers of tweets are counted for 28 days. Unquarantined users’ total and average tweeted numbers are displayed using dashed lines in blue and purple, respectively.

#### Changing patterns of mental status

Figure 2 demonstrates the changes in anxious depression (AD) level of quarantined users before, during, and after quarantine and the 28-day AD level change of unquarantined users. The data of both groups was collected in the same window to ensure a similar background of potentially AD-stimulating news and trends. The AD level change range of the unquarantined group is slightly smaller than that of the quarantined group, ranging between 0.119 and 0.154 compared to 0.138 and 0.166, respectively, across the four weeks under investigation. There are slight changes across the 28-day for unquarantined users with no obvious specific feature as compared to quarantined users. Days 1 and 13, the start and end dates of the 2-week quarantine, are clearly AD inflection points. The AD level of quarantined users increased at Day 1 of quarantine, followed a clear descending trend as the quarantine progressed, and declined rapidly when the quarantine was about to end. On Day 13, one day before the end of quarantine, AD dropped to 0.118. By that date, the clinical features of COVID-19 infection, including fever, malaise, dry cough, and shortness of breath should have surfaced if the person was infected^28^ except in rare cases of more than 14 days’ incubation or where the individual was asymptomatic. After Day 13, quarantined users tweeted regularly (see Figure 1), but their AD level dropped steadily. After their quarantine, they are relieved.

**Figure 2:**
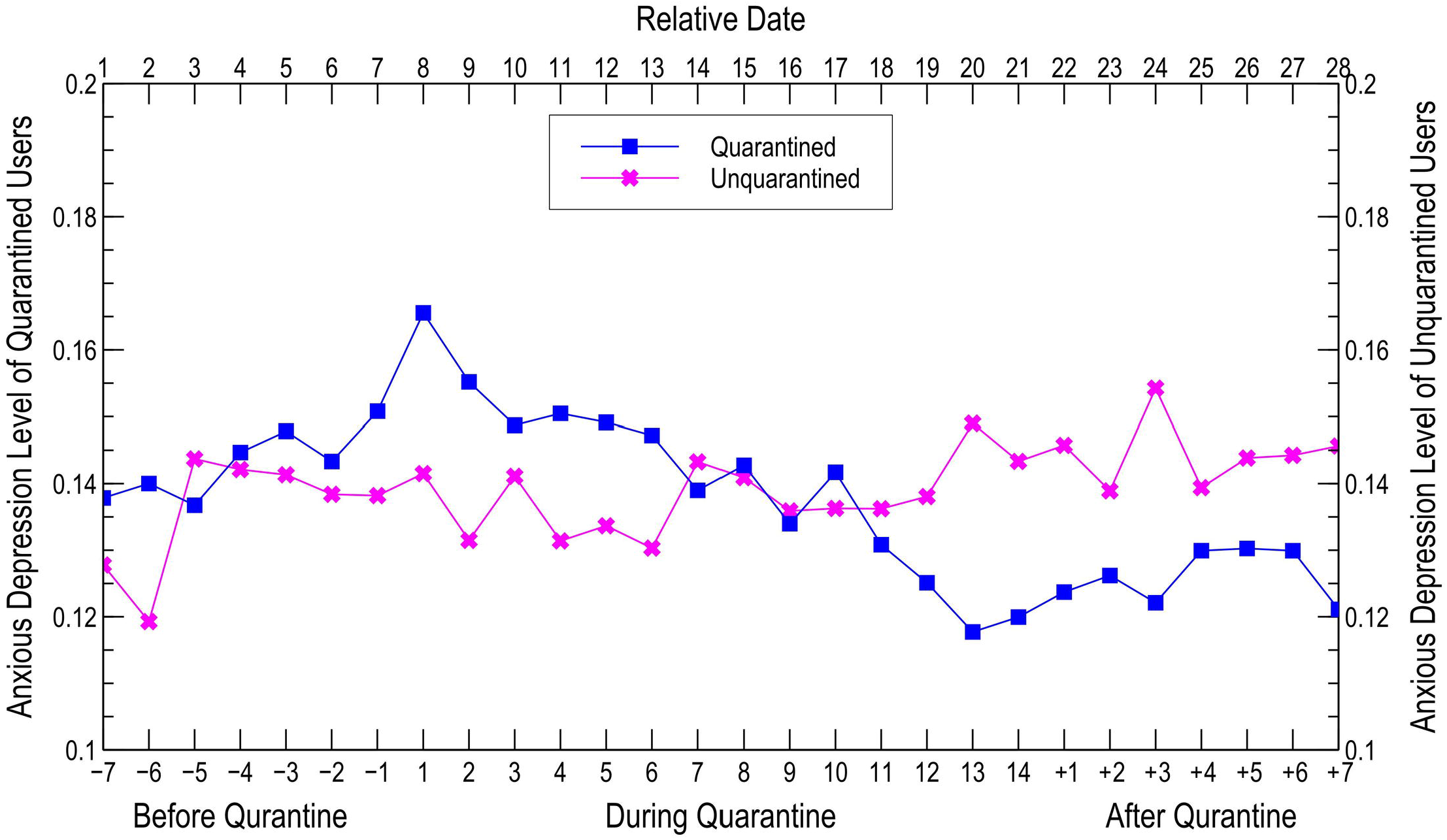
Anxious depression level comparison between quarantined and unquarantined users. The anxious depression level of a certain day is the ratio of the number of tweets containing the keywords from the lexicon to the number of total tweets on that day.

Figure 3 displays the top six keywords appearing in the tweets of all investigated users. Nine keywords appear in the displayed lists of both quarantined and unquarantined users. They are “bad”, “death”, “die”, “down”, “mean”, “sick”, “symptoms”, “virus”, and “wrong”. This suggests both groups felt bad / down about COVID-19 deaths and were afraid of symptoms. During the quarantine period, the keywords “isolation”, “down”, “virus”, and “bad” frequently appeared in quarantined users’ tweets. They also tended to swear a lot, which we interpreted as a sign of anxiety,^29^ and they mentioned “symptoms” and “die” frequently, revealing fears of infection and death. A closer look at users’ tweets on Day 1 of quarantine shows that they tweeted more about “isolation”, “virus”, and “down” than on any other day in the 28-day period. “Sick”, “symptoms”, and even “die” came into their tweets on many days. For unquarantined users, “die” and “virus” were the top two keywords in everyday most frequently used words, while “down” and “death” are the third and fourth most frequently used. This implies that they were worried about the virus itself and the death it caused, whereas quarantined users worried about themselves.

**Figure 3:**
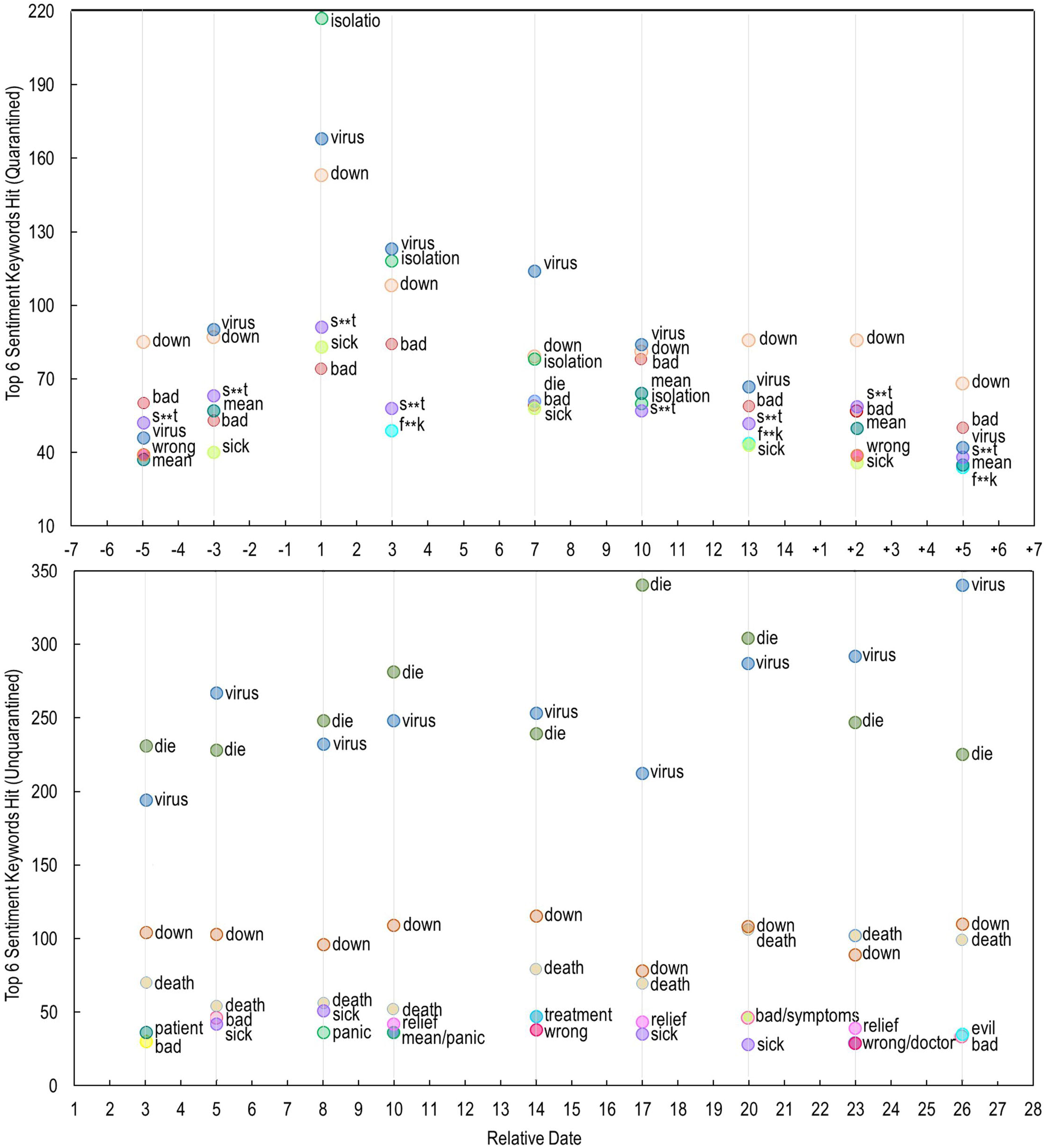
Top six anxious depression keywords in tweets by quarantined and unquarantined users. The daily top six keywords are the most frequently appearing keywords from the anxious depression lexicon. Figure 3 displays keywords only on specific days. A total of 22 keywords are most frequently used.

It is surprising to see the high levels of anxious depression among quarantined users in the 7 days before quarantine. On average, mental states were worse at this stage than during or after quarantine. This well reflected the panic of the public, which had very limited knowledge of this COVID-19 virus when it was outbreak.

A wary trend is the resurgence of anxious depression after the 2-week quarantine period. Figure 3 shows anxious depression levels rising from the lowest point of 0.118 on Day 13 during quarantine to around 0.130 in Days 4, 5, and 6 after quarantine. We attribute this to the fact that most people were still locked down at home due to social distancing requirements^30^. After a 14-day quarantine, one must become worn and irritated because of boredom, frustration, and possibly financial loss, although the alarm of fearing infection can be disarmed. During this period, post-quarantine users still swore a lot and their tweets made frequent use of the words “down”, “bad”, “virus”, and the like.

## Discussion

This research conducts a longitudinal analysis of the psychological states before, during, and after quarantine. Our analyses show that quarantine has non-negligible negative psychological impacts on those who are quarantined. In fact, these impacts took effect even before formal quarantine began. At this early stage, our knowledge about the virus with regards to its basic reproduction number (R0), mode of transmission, incubation period, and symptoms is rather limited. Governments inevitably showed hesitation to adopt the most effective containing strategies. This is further exacerbated by the fact that we still do not have vaccines to prevent infection or drugs to cure the infected. To mitigate the negative psychological impacts, governments or their disease control and prevention (CDC) centers should release authoritative information, telling people what is happening and why, and explaining how long it will continue^2^.

We discovered that among quarantined individuals, anxious depression peaked on Day 1 of quarantine, remained at a high level during the first week of quarantine, and then declined sharply to the end of the quarantine. This tallies with the medical finding that the COVID-19 incubation period ranges from 1 to 14 days, and is most commonly 5 days.^31^ After 5 days most quarantined individuals should have seen the symptoms if they were infected and would perhaps be hospitalized. Otherwise, they can be much relieved. During this period, quarantined people need good communication and authoritative information, e.g., about the death rates, availability of medical resources, or vaccine or drug therapies developing progress. They need care, encouragement, and emotion management. Our human communities have done many noble things, e.g., Queen Elizabeth’s national address broadcasting to the whole world, entertainment companies opening their resources for free, the “One World: Together at Home” concert curated by Lady Gaga, or the like.

A particularly meaningful finding of this research is that there is a post-quarantine resurgence in anxious depression. Unlike in previous studies of PTSD after quarantine, the sample under our investigation was still lock down indoor. While relieved of fear of infection, new psychological impacts were witnessed. These were probably caused by boredom, loneliness, being cooped up with family members, or financial concerns. While the above mentioned caring, encouragement, and emotion management measures can still be promoted, a more proactive strategy is to rethink the implementation of a uniform lockdown policy, which is mechanically extended again and again in most of the countries. Some carefully thought-out strategies to unlock the cities should be devised to alleviate the negative impacts of quarantine lockdown.

## Conclusions

This research investigates the adverse psychological impacts of COVID-19 quarantine. It was found that quarantined people experienced a clear pattern of change in mental state before, during, and after quarantine. They suffered a high level of anxious depression before quarantine, probably owing to infection fears, confusion, and misleading information, all of which can be exacerbated by the mass and social media. The anxious depression level spiked on Day 1 of quarantine but gradually diminished, reaching its lowest point on Day 13. After two weeks’ quarantine, most clinical features of COVID-19 should have surfaced if an infection were present, meaning that fear of infection can subside. Nevertheless, the anxious depression level had a resurgence post quarantine, probably because most subjects were still in lockdown in accordance with local regulations. The contributions of this paper are twofold: (a) evidence on the psychological effects before, during, and after quarantine during the COVID-19 outbreak; and (b) a straightforward but innovative sentimental analysis approach to access the subjects when the traumatic event is still going on.

Whilst the necessity and effectiveness of quarantine in containing the COVID-19 pandemic are evident, its negative psychological impacts cannot be neglected. It takes effects even before the quarantine is formally started. Our research provides evidence backing up calls for officials to ensure the quarantine experience is as tolerable as possible. Measures such as authoritative information, caring, entertaining, encouragement, emotion management, and the like should be consciously devised to alleviate the negative psychological impacts. Our research also provides evidence in support of calls to keep quarantine lockdowns as short as possible, as negative psychological impacts will return if the post-quarantine individual is further grounded indoors. Therefore, delicate strategies to unlock the cities, instead of implementing a uniform and brutal lockdown policy, are highly desired.

## Data Availability

All the data for this research is collected from Twitter.

https://github.com/EaGlEtSuImars/The-Psychological-effects-of-quarantine-during-COVID.git.

## Acknowledgments

We acknowledge the Tweeter for making the tweets available for researchers. Author contributions: WL, conceived the study. LY. curated data. LY. and JX. performed the analysis. WL, JX, and LY. wrote the first draft of the manuscript. WL, BZ, TL, and CW reviewed and edited the manuscript. Competing interests: All authors declare no competing interests. Data and materials availability: All code and data are available in the supplementary materials.

## Supporting information captions

Table S1. Lexicons of anxious depression

Data and code URL

## Notes

### Competing Interest Statement

The authors have declared no competing interest.

### Funding Statement

No funding support for this study.

### Author Declarations

Prof. T.S. Veitch, Chairman, Human Resource Ethics Committee, The University of Hong Kong

## References

1. Leleu H, Hoertel N, Massetti M, et al. Should we quickly get out of confinement to face COVID-19 pandemic? Estimating the medical outcomes of different public health strategies using a robust microsimulation agent-based model. 2020. Preprint: doi:10.2139/ssrn.3569844.

2. Brooks SK, Webster RK, Smith LE, et al. The psychological impact of quarantine and how to reduce it: Rapid review of the evidence. Lancet. 2020; 395:912–920.

3. Manuell ME, Cukor J. Mother Nature versus human nature: Public compliance with evacuation and quarantine. Disasters. 2011; 35:417–442.

4. Van der Kolk BA, McFarlane AC, Eds., Traumatic stress: The effects of overwhelming experience on mind, body, and society. New York, NY: Guilford Press; 1996.

5. Hawryluck L, Gold WL, Robinson S, et al. SARS control and psychological effects of quarantine, Toronto, Canada. Emerging Infectious Disease. 2004; 10:1206–1212.

6. Liu X, Kakade M, Fuller CJ, et al. Depression after exposure to stressful events: Lessons learned from the severe acute respiratory syndrome epidemic. Comprehensive Psychiatry. 2012; 53:15–23.

7. Derluyn I, Broekaert E, Schuyten G, et al. Post-traumatic stress in former Ugandan child soldiers. Lancet. 2004; 363:861–863.

8. Spitzer RL, Kroenke K, Williams JB, et al. A brief measure for assessing generalized anxiety disorder: The GAD-7. Archives of Internal Medicine. 2006; 166:1092–1097.

9. Lievaart M, Franken IH, Hovens JE. Anger assessment in clinical and nonclinical populations: Further validation of the State–Trait Anger Expression Inventory-2. Journal of Clinical Psychology. 2016; 72:263–278.

10. Schaufeli WB, Salanova M, González-Romá V, et al. The measurement of engagement and burnout: A two sample confirmatory factor analytic approach. Journal of Happiness Studies. 2002; 3:71–92.

11. Taylor MR, Agho KE, Stevens GJ, et al. Factors influencing psychological distress during a disease epidemic: Data from Australia’s first outbreak of equine influenza. BMC Public Health. 2008; 8:347.

12. Saha K, Chan L, De Barbaro K, et al. Inferring mood instability on social media by leveraging ecological momentary assessments. In: Proceedings of the ACM on Interactive, Mobile, Wearable and Ubiquitous Technologies. 2017; 1–27.

13. The Washington Post. Our iPhone weekly screen time reports are through the roof, and people are ‘horrified’. 2020. Available at: https://www.washingtonpost.com/technology/2020/03/24/screen-time-iphone-coronavirus-quarantine-covid/. Accessed April 18, 2020.

14. Kumar A, Sharma A, Aror A. Anxious depression prediction in real-time social data. In: International Conference on Advances in Engineering Science Management & Technology 2019; 1–7.

15. Boiy E, Hens P, Deschacht K, et al. Automatic sentiment analysis of on-line text. In Proceedings of the 11th International Conference on Electronic Publishing. 2007;349–360.

16. Coppersmith G, Harman C, Dredze M. Measuring post-traumatic stress disorder in Twitter. In Eighth International AAAI Conference on Weblogs and Social Media. 2014; 579–582.

17. De Choudhury M, Counts S, Horvitz EJ, et al. Characterizing and predicting postpartum depression from shared Facebook data. In Proceedings of the 17th ACM Conference on Computer Supported Cooperative Work & Social Computing. 2014; 626–638.

18. Pedersen T. Screening Twitter users for depression and PTSD with lexical decision lists. In Proceedings of the 2nd Workshop on Computational Linguistics and Clinical Psychology: From Linguistic Signal to Clinical Reality. 2015; 46–53.

19. De Choudhury M, Counts S, Horvitz EJ. Social media as a measurement tool of depression in populations. In Proceedings of the 5th Annual ACM Web Science Conference. 2013; 47–56.

20. Sasaki T, Akaho R, Sakamaki H, et al. Mental disturbances during isolation in bone marrow transplant patients with leukemia. Bone Marrow Transplantation. 2000; 25:315–318.

21. Zimmerman M, Kerr S, Kiefer R, et al. What is anxious depression? Overlap and agreement between different definitions. Journal of Psychiatric Research. 2019; 109:133–138.

22. Ionescu DF, Niciu MJ, Henter ID, et al. Defining anxious depression: A review of the literature. CNS Spectrums. 2013; 18:252–260.

23. WHO. Depression. 2020. Available at: https://www.who.int/news-room/fact-sheets/detail/depression. Accessed April 12, 2020.

24. Evans DL, Foa EB, Gur RE, et al., Eds. Treating and preventing adolescent mental health disorders: What we know and what we don’t know. Oxford, Oxford University Press; 2005.

25. Lambert AJ, Eadeh FR, Peak SA, et al. Toward a greater understanding of the emotional dynamics of the mortality salience manipulation: Revisiting the “affect-free” claim of terror management research. Journal of Personality and Social Psychology. 2014; 106:655–678.

26. Hunter R, Gough A, O’Kane N, et al. Ethical issues in social media research for public health. American Journal of Public Health. 2018; 108:343–348.

27. Lomborg S, Bechmann A. Using APIs for data collection on social media. The Information Society. 2014; 30:256–265.

28. Wang Y, Wang Y, Chen Y, et al. Unique epidemiological and clinical features of the emerging 2019 novel coronavirus pneumonia (COVID-19) implicate special control measures. Journal of Medical Virology. 2020; 92:568–576.

29. Shen JH, Rudzicz F. Detecting anxiety through Reddit. In Proceedings of the Fourth Workshop on Computational Linguistics and Clinical Psychology—From Linguistic Signal to Clinical Reality. 2017; 58–65.

30. Hale T, Petherick A, Phillips T, et al. Variation in government responses to COVID-19: Version 4.0. (Working Paper. Blavatnik School of Government, Oxford University, 2020). https://www.bsg.ox.ac.uk/sites/default/files/2020-04/BSG-WP-2020-031-v4.0_0.pdf

31. Lauer SA, Grantz KH, Bi Q, et al. The incubation period of coronavirus disease 2019 (COVID-19) from publicly reported confirmed cases: Estimation and application. Annals of Internal Medicine. 2020; 172(9):577–582.

